# Hepatoprotective Efficacy of GeneIII^®^ L-Ergothioneine Capsules: A Self-Controlled Clinical Trial

**DOI:** 10.64898/2025.12.30.25343022

**Authors:** Rujun He, Wei Ding, Juan Cao, Hongying Ju, Fengjuan Liu, Guohua Xiao

## Abstract

**Context:** Erogothioniene (EGT), a potent natural antioxidant, exerts hepatoprotection. However, human clinical evidence remains absent.

**Objective:** to observe the clinical efficacy and safety studies of GeneIII^®^ L-Ergothioneine Capsules in the Hepatoprotective Efficacy.

**Materials and methods:** Thirty subjects with some abnormal liver function indicators were selected to take 2 capsules of 30mg GeneIII^®^ L-Ergothioneine Capsules per day, for 30 consecutive days. The changes in serum liver function biomarkers, clinical efficacy self-rating scale, and Pittsburgh sleep quality index scale (PSQI) from baseline in different visit cycles were compared by self-control. All adverse events that occured after receiving the trial medication were also tracked and the incidence of adverse events during the trial period was calculated.

**Results:** All subjects completed treatment and follow-up. After taking GeneIII^®^ L-Ergothioneine Capsules, the subjects’ aspartate aminotransferase (AST) levels were significantly reduced compared to baseline (p=0.0082). Additionally, alanine aminotransferase (ALT) levels were also significantly reduced (p=0.0025), gamma-glutamyl transferase (GGT) levels were significantly reduced compared to baseline (p=0.0270). Physical function scores on the Clinical Efficacy Self-Assessment Scale improved significantly compared to baseline. Physical function scores decreased significantly by 39.04% compared to baseline (p<0.0001). Additionally, the level of daytime functional impairment on the Pittsburgh Sleep Quality Index was significantly reduced by 51.56% compared to baseline (p=0.0038). No adverse events occurred throughout the study. No side effects were observed clinically, confirming the reliable safety of GeneIII^®^ L-Ergothioneine Capsules under the current study design.

**Conclusion:** GeneIII^®^ L-Ergothioneine Capsules have a significant effect on improving liver function, physical function, and sleep quality in subjects, and the overall safety is good.

## Introduction

The liver serves as the central organ responsible for metabolism, detoxification, and synthesis in the human body, overseeing the metabolism of macromolecules such as fats, proteins, and carbohydrates. Its health status directly affects overall physiological homeostasis. The liver is highly susceptible to damage from various toxic substances, and the global incidence of chronic liver diseases (such as fatty liver disease, viral hepatitis, drug-induced liver injury, and cirrhosis) continues to rise [Chan, B. K. Y et al. 2021]. The liver is a particularly vulnerable organ to reactive oxygen species (ROS), which are not only produced during metabolic processes but also during the biotransformation of exogenous compounds. Disruption of redox balance leads to oxidative stress, and ROS contributes to the progression of various liver diseases by inducing liver cell damage, apoptosis, and haematopoietic stem cell activation, thereby impairing liver function and causing disease [Abdolamir Allameh et al. 2023].

Each hepatocyte contains 1,000–2,000 mitochondria, which play a role in maintaining the redox state of the cell by balancing the production of reactive oxygen species (ROS) and the clearance of ROS by the antioxidant defence system. When damaged mitochondria are unable to clear excess ROS, oxidative stress occurs, which is considered to be one of the factors leading to hepatocyte death and liver damage [Ping Chen et al.2024].

Therefore, it is scientifically valuable to identify natural functional substances with strong antioxidant properties that can enter mitochondria to clear ROS and alleviate liver damage caused by oxidative stress.

Ergothioneine (EGT) is a thio-histidine bataine amino acid, which was discovered and isolated by Charles Tanley in 1909 during his research on ergot fungus (Claviceps purpurea) [Tanret C et al. 1909]. EGT is an endogenous amino acid that cannot be synthesised by the human body and must be obtained through diet. One of its advantages is its role as an endogenous antioxidant, efficiently scavenging hydroxyl radicals, chelating with ferrous and copper ions, inhibiting the production of hydroxyl radicals by H_2_O_2_, and activating the cell’s natural antioxidant defence mechanisms. Additionally, it can be absorbed by human cells via the specific transporter SLC22A4 (OCTN1) and accumulates in various tissues such as the liver, eye, blood, kidneys, spleen, and lungs. EGT was described as a “longevity vitamin”and exploratory research has proposed an association between EGT and reduced pathology in conditions [Bindu D. Paul 2022]. Recent research shows that EGT can be a biomarker of cognitive reserve against amyloid pathology, suggests potential beneficial effects of EGT metabolism [Joyce R. Chong et al. 2025]. EGT may be useful in the prevention and/or treatment of Parkinson’s disease (PD) [Roda, E et al. 2023; Teruya, T et al. 2021].besides, EGT has been proven to be effective in delaying or preventing cataracts[Shukla, Y., et al 1981], improving sleep[Matsuda, Y te al. 2020; Makoto Katsube a et al. 2022 ], improving cardiovascular function[Smith E et al. 2020 ], and treating diabetes[Dare, A., ert al. 2021] and syndrome (ErgMS)[Tian, X et al. 2021].

Studies have shown that EGT can be accumulated in the liver in a guinea pig model of non-alcoholic fatty liver disease [Shelly Lu et al. 2008]. Many studies have demonstrated that it has a promising effect on the improvement of liver function.

Evidence suggests that liver damage can be assessed by establishing a CCl_4_-induced liver injury model and measuring serum transaminase activity and liver antioxidant enzyme levels. liver injury was effectively prevented in ergothioneine-treated mice [Xiong et al.2024]. Supplementation with L-ergothioneine not only protects the liver from lipid peroxidation damage but also reduces the consumption of endogenous glutathione and α-tocopherol [Saravanabhavan S et al.2022; Monica D et al. 2004; Chieko K et al.2005]. EGT-induced upregulation of hsp70 also protects the liver from ischaemia-reperfusion injury, thereby reducing lipid peroxidation [Abdulkadir B et al.2004]. EGT may inhibit glycerophospholipid metabolism via PLA2G2A, thereby suppressing the TGF- β/Smads signalling pathway and alleviating liver fibrosis. EGT may be an effective therapeutic strategy for reversing liver fibrosis [Alfonsina M et al. 2019; Mao et al.2025]. These findings suggest that ergothioneine is an ideal candidate substance for improving liver function. Currently, there are no human clinical studies on the effects of ergothioneine on liver function improvement. This study aims to investigate the efficacy and safety of GeneIII^®^ L-Ergothioneine Capsules in improving liver function through a human self-control study.

## Materials and Methods

Prospective study. A single-centre, randomised, open-label, parallel, self-controlled clinical trial was conducted, enrolling eligible subjects from December 2024 to January 2025 at the Qingdao Central Hospital of Rehabilitation University. The study comprised a screening period, enrolment period, trial period, and follow-up period. Efficacy assessment was conducted at the end of the study, and safety assessment was performed throughout the entire study process.

This study enrolled a total of 30 subjects, with no gender restrictions. Subjects were instructed to take 2 Gene III^®^ L-Ergothioneine Capsules (30 mg EGT per capsule) of ergothioneine daily, either as a single dose or divided into two doses (1 capsule in the morning and 1 capsule in the evening), for a continuous period of 30 days. All GeneIII^®^ L-Ergothioneine Capsules were provided by Gene III Biotechnology Co., Ltd. (main ingredients: ergothioneine, maltodextrin, hydroxypropyl methylcellulose, magnesium stearate, etc.). This study strictly adhered to the ethical guidelines of the Declaration of Helsinki. The trial protocol was reviewed and approved by an Independent Ethics Committee (IEC), and the entire ethical review process complied with Good Clinical Practice (GCP), the Declaration of Helsinki, and relevant domestic laws and regulations. The study protocol was reviewed by the Ethics Review Committee of Qingdao Central Hospital of Rehabilitation University (Ethics Number:KY202416002). All subjects signed written informed consent forms prior to participating in the study to ensure their legal rights and interests.

Inclusion criteria: All inclusion criteria must be met to be eligible for inclusion. (1) Age 18–65 years, no gender restrictions; (2) Willingness to comply with the doctor’s intervention plan and cooperate with follow-up, informed consent, and signing of the informed consent form; (3) Regular lifestyle and eating habits, stable weight over the past three months; (4) Abnormal levels of any two of the following serum enzyme markers: aspartate aminotransferase (AST), alanine aminotransferase (ALT), gamma-glutamyl transferase (GGT), or alkaline phosphatase (ALP). AST abnormal range: >40 U/L; ALT abnormal range: >40 U/L for males, >35 U/L for females; GGT abnormal range: >50 U/L; ALP abnormal range: >110 U/L; (5) After assessment of the subject’s condition and relevant indicators by a gastroenterologist, including aspartate aminotransferase (AST), alanine aminotransferase (ALT) levels, gamma-glutamyl transferase (GGT), and alkaline phosphatase (ALP), the gastroenterologist determines whether the subject is suitable for participation in this project.

Exclusion criteria: Meeting any one exclusion criterion results in exclusion. (1) Subjects with severe concomitant diseases such as heart, liver (viral hepatitis, cirrhosis), kidney, or haematopoietic system disorders, or psychiatric disorders; (2) Subjects who have taken medications or supplements known to affect fatty liver or body fat within the past month; (3) Subjects allergic to the investigational product; (4) Pregnant women, women planning to become pregnant, or breastfeeding women; (5) Subjects with elevated ALP levels due to skeletal diseases, rickets, osteomalacia, fibrocystic osteitis, osteoporosis, bone tumours, fractures in the healing phase, hyperparathyroidism, rheumatoid arthritis, or dysplastic osteitis; (6) Subjects unable to abstain from alcohol during the trial period; (7) Subjects with clinically significant abnormalities in vital signs monitoring, physical examination, clinical laboratory tests (complete blood count, urinalysis), urine pregnancy test (female only), or viral hepatitis testing; (8) Subjects who have participated in or are currently participating in other clinical studies within the past three months; (9) Subjects deemed by the investigator to be unsuitable for this trial or at risk of loss to follow-up.

The treatment cycle for this trial is one course of treatment with this product. The dosage is 30 mg EGT per capsule, taken twice daily. The product should be taken twice daily, either as a single dose or one capsule in the morning and one in the evening, for 30 consecutive days.A total of 30 subjects were enrolled in this trial, all of whom took ergothioneine capsules. Three visits were conducted: prior to product administration (baseline period), 15 days after product administration (trial period), and 30 days after product administration (follow-up period). The assessment criteria included liver serum markers, clinical efficacy self-assessment scales, the Pittsburgh Sleep Quality Index (PSQI), and changes from baseline. Each visit included testing of the aforementioned indicators and completion of questionnaires. The study compared changes in seliver serum markers, clinical efficacy self-assessment scales, Pittsburgh Sleep Quality Index, and adverse event incidence rates between different visit periods and baseline.

## Statistical analysis

efficacy evaluation indicators and Safety evaluation indicator were analyzed, Statistical tests were performed using SAS 9.4 software. Paired t-tests were used to compare differences between different visits after product use and baseline. If the data did not meet the assumptions of normal distribution or homogeneity of variance, rank-sum tests were used instead. A paired t-test was used to compare differences between different product specifications at the same visit after product use. If the data did not meet the assumptions of normal distribution or homogeneity of variances, a rank-sum test was used instead. The significance level was set at α= 0.05.Statistical analysis of safety evaluation indicators was performed using SAS 9.4 software. Adverse events occurring in this study were described according to the type of product used, and the incidence rate, correlation with the study product, and severity of adverse events were statistically described.

## Results

### Demographic and baseline characteristics

This study enrolled a total of 30 subjects, with no gender restrictions. All the subjects completed the trial (figure 1). Subjects were of an average age of 43.37±9.94 years (range:27-62).18 (60%) subjects were male.

**Figure 1.**
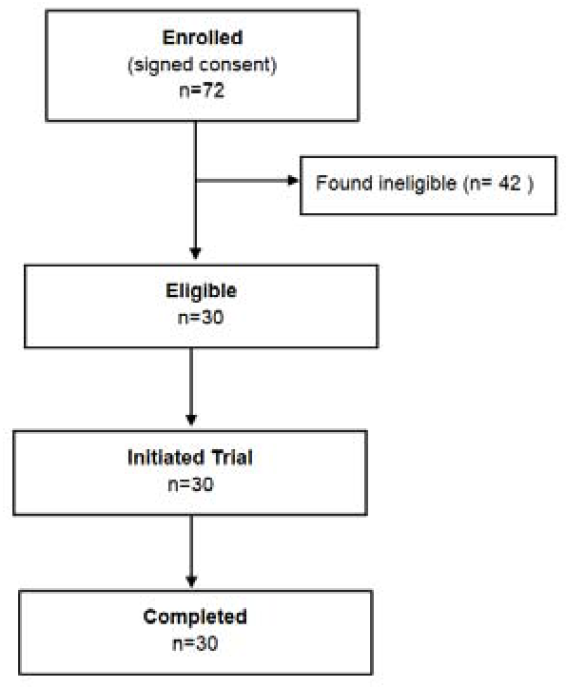
participant flow chart. Flow chart of participants enrolled in the trial.

### Clinical response

The assessment criteria included liver serum markers, clinical efficacy self-assessment scales, the Pittsburgh Sleep Quality Index (PSQI), and changes from baseline.

After taking GeneIII^®^ L-Ergothioneine Capsules for 15 and 30 days, the subjects’ aspartate aminotransferase (AST) levels were significantly lower than at baseline. The average AST level at baseline was 47.5 U/L; after 15 days of administration, the average level was 41.5 U/L, representing a significant decrease of 12.69% compared to baseline (p=0.0350); After 30 days of administration, the average level was 38.2 U/L, representing a significant reduction of 19.64% compared to the baseline period (p=0.0082), as shown in Figure 2. After taking GeneIII^®^ L-Ergothioneine Capsules for 15 and 30 days, the subjects’ alanine aminotransferase (ALT) levels decreased significantly. The baseline average level of alanine aminotransferase (ALT) was 80.9 U/L; after 15 days of administration, the average level was 68.3 U/L, representing a significant decrease of 15.57% compared to the baseline (p=0.0029); After 30 days of administration, the average level was 63.7 U/L, representing a significant decrease of 21.25% compared to the baseline (p=0.0025).

**Figure 2.**
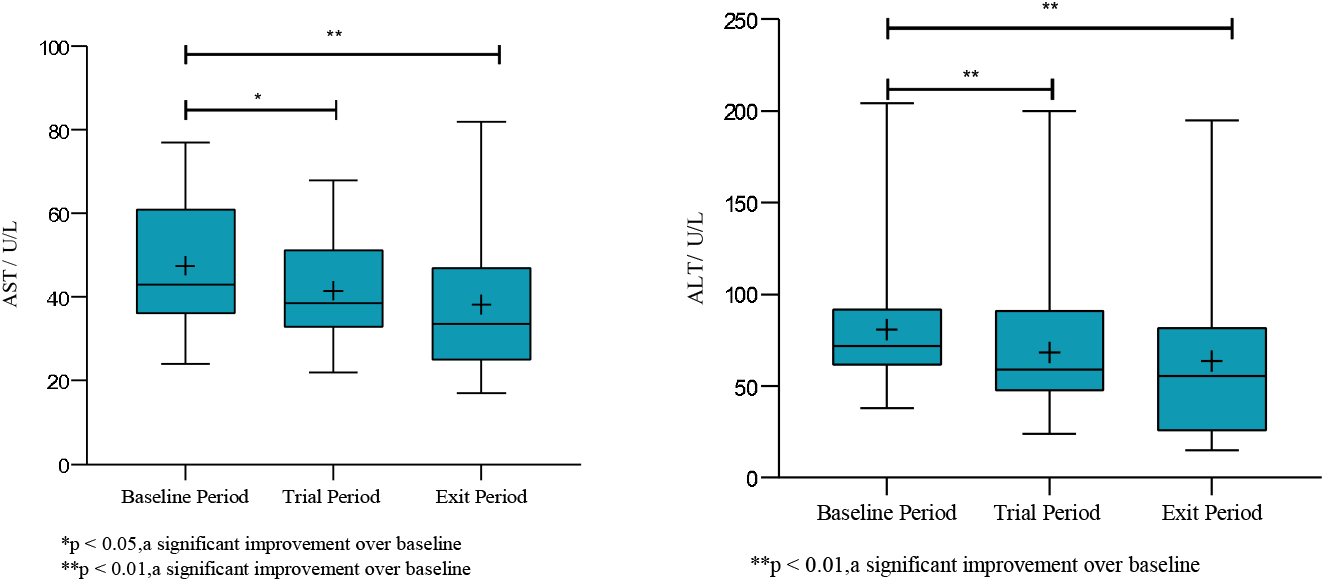
Box plot of aspartate aminotransferase (AST) and alanine aminotransferase (ALT) levels 15 and 30 days after taking the product

After taking GeneIII^®^ L-Ergothioneine Capsules for 15 days, the subjects’ gamma-glutamyltransferase (GGT) levels decreased significantly. The baseline average level of gamma-glutamyltransferase (GGT) was 87.1 U/L; after 15 days of administration, the average level was 77.1 U/L, representing a significant decrease of 11.49% compared to the baseline (p=0.0270); after 30 days of administration, the average level was 71.2 U/L, as shown in Figure 3.

**Figure 3.**
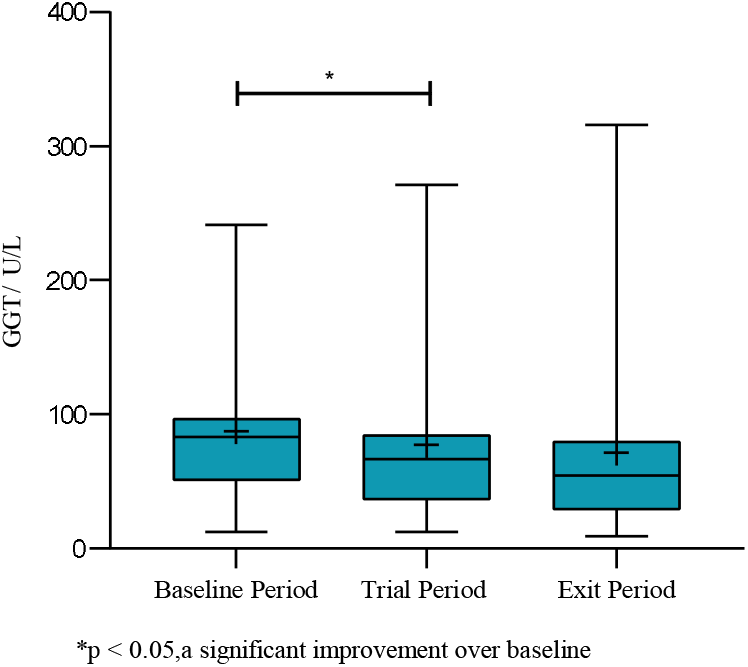
Box plot of gamma-glutamyltransferase (GGT) levels 15 and 30 days after taking the product

**Figure 4.**
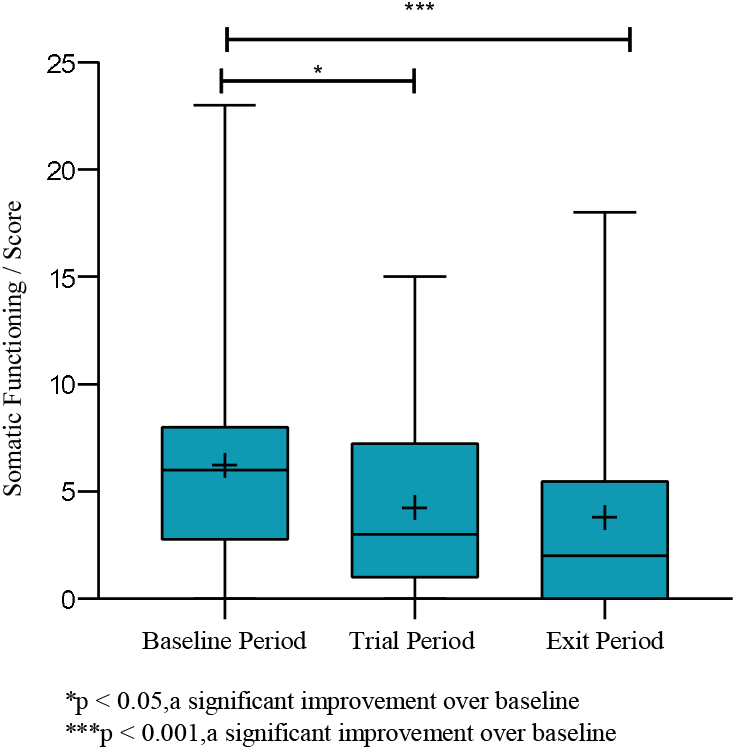
Box plot of physical function levels 15 and 30 days after taking the product

The physical function level in the Clinical Efficacy Self-Assessment Scale is a comprehensive assessment of the subject’s subjective fatigue, fatigue recovery status, dizziness, shortness of breath, sleep issues, discomfort in the liver area, and dry eyes. After taking GeneIII^®^ L-Ergothioneine Capsules for 15 and 30 days, the subjects’ physical function levels improved significantly compared to the baseline period. The average physical function level at baseline was 6.2 points; after 15 days of administration, the average level was 4.3 points, representing a significant decrease of 31.55% compared to baseline (p=0.0385); after 30 days of administration, the average level was 3.8 points, representing a significant decrease of 39.04% compared to baseline (p<0.0001), as shown in Figure 5.

**Figure 5.**
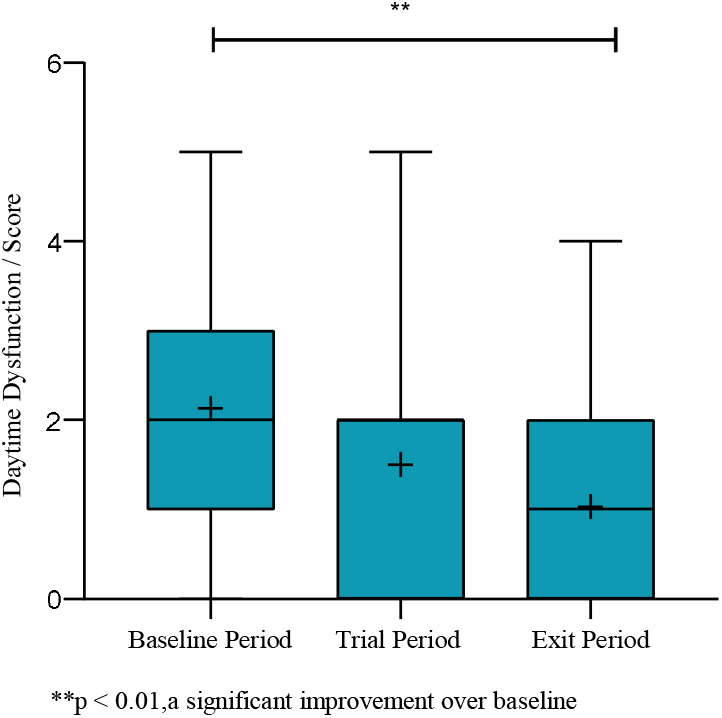
Box plot of daytime functional impairment levels 15 and 30 days after taking the product

The Pittsburgh Sleep Quality Index (PSQI) assesses daytime functional impairment based on participants’ perceived sleepiness and energy levels. After taking GeneIII^®^ L-Ergothioneine Capsules for 30 days, participants’ daytime functional impairment levels improved significantly compared to baseline. The average level of daytime dysfunction at baseline was 2.1 points, and after 30 days of administration, the average level was 1.0 points, representing a significant decrease of 51.56% compared to the baseline period (p=0.0038), as shown in Figure 5.

Thirty subjects were enrolled in this trial. No adverse events occurred in any of the subjects, and there was no impact on changes in liver synthetic function or lipid levels. No other side effects (safety indicators) were observed clinically, confirming that ergothioneine has reliable safety under the current study design.

## Discussion

Extensive research has confirmed that the absorption and transport of ergothioneine depend on the specific organic cation transporter encoded by the SLC22A4 gene (organic cation transporters novel-1, OCTN-1). The widespread expression of OCTN-1 ensures that ergothioneine is distributed throughout all organs of the body, including the liver, which is susceptible to oxidative damage. Mitochondria are the primary sites for free radical generation within cells. Ergothioneine enters mitochondria via OCTN-1, effectively neutralising free radicals and reducing mitochondrial oxidative damage. Numerous studies have explored the role of antioxidant pathways in improving liver function. As a cellular mitochondrial-level antioxidant, ergothioneine demonstrates significant potential in protecting liver function. According to a study by Palma Ann Marone et al. [Marone, P. A et al., 2016], rats underwent a 90-day repeated-dose oral administration study with dose levels of 0, 400, 800, and 1600 mg/kg body weight/day, and no adverse effects were observed in clinical parameters, body weight/weight gain, food consumption and efficiency, clinical pathology, or histopathology. According to the EFSA [Turck, D et al., 2017], the NOAEL for ergothioneine is 800 mg/kg body weight/day, indicating extremely high safety.

The results of this study indicate that oral administration of GeneIII^®^ L-Ergothioneine Capsules for 15 and 30 days can effectively reduce liver function indicators AST, ALT, and GGT levels, with significant reductions in these indicators compared to baseline, thereby effectively improving liver function. Additionally, the daytime dysfunction level on the Pittsburgh Sleep Quality Index was significantly reduced compared to baseline. A total of 30 subjects were included in the safety data set, with no adverse events reported. The overall safety profile of the study product was good.

## Conclusions

In summary, oral GeneIII^®^ L-Ergothioneine Capsules can effectively improve liver function indicators and demonstrate good overall safety. This study still has certain limitations: (1) This study used a self-controlled design, which although helps eliminate the influence of individual differences, cannot rule out time effects and regression effects. Future studies should conduct placebo-controlled or positive-controlled trials to more accurately assess treatment efficacy; (2) The sample size in this study was relatively small, which to some extent limits the statistical power and generalisability of the results. Future studies should expand the sample size to further enhance the generalisability and reliability of the research findings;

## Data Availability

All data produced in the present work are contained in the manuscript

## Data Availability Statement

Data will be made available from the corresponding author upon reasonable request.

## Acknowledgements

We would like to acknowledge Gene III Biotechnology Co., Ltd. for providing the research funding. Gratitude also goes to Anhui Wanbang Pharmaceutical Technology Co., Ltd. for resource docking, and to Qingdao Central Hospital for providing support in clinical trials. Many thanks to Professor Xuefeng Zhou for his support of the clinical project.

## References

Chan, B. K. Y., Elmasry, M., Forootan, S. S., Russomanno, G., Bunday, T. M., Zhang, F., & Copple, I. M. (2021). Pharmacological activation of Nrf2 enhances functional liver regeneration. Hepatology, 74(2), 973–986.

Allameh, Abdolamir, Reyhaneh Niayesh-Mehr, Azadeh Aliarab, Giada Sebastiani, and Kostas Pantopoulos. 2023. “Oxidative Stress in Liver Pathophysiology and Disease” Antioxidants 12, no. 9: 1653.

Chen P, Yao L, Yuan M, Wang Z, Zhang Q, Jiang Y, Li L (2023). Mitochondrial dysfunction: A promising therapeutic target for liver diseases. Genes Dis. 11(3).

Tanret C (1909). Sur une base nouvelle retiree du seigle ergote,l’ergothioneine. Compt Rend. 149:222–4.

Paul B. D. (2022). Ergothioneine: A Stress Vitamin with Antiaging, Vascular, and Neuroprotective Roles?. Antioxidants & redox signaling, 36(16-18), 1306–1317.

Joyce R. Chong, Irwin K. Cheah, Richard M. Tang, Barry Halliwell, Christopher P. Chen, Mitchell K.P. Lai. (2025). Metabolism of the antioxidant Ergothioneine as a Plasma Biomarker of Cognitive Reserve. the preprint server for health sciences.

Roda, E., De Luca, F., Ratto, D., Priori, E. C., Savino, E., Bottone, M. G., & Rossi, P. (2023). Cognitive Healthy Aging in Mice: Boosting Memory by an Ergothioneine-Rich Hericium erinaceus Primordium Extract. Biology, 12(2), 196.

Teruya, T., Chen, Y. J., Kondoh, H., Fukuji, Y., & Yanagida, M. (2021). Whole-blood metabolomics of dementia patients reveal classes of disease-linked metabolites. Proceedings of the National Academy of Sciences of the United States of America, 118(37), e2022857118.

Tng, T. J. W., Leow, D. M. K., Goh, G., Wang, Z., Liu, Y., Tang, R. M. Y., Lai, K. C. L., Basil, A. H., Choong, H. C., Goh, W. W. B., Cheah, I. K., Ong, W. Y., Halliwell, B., & Lim, K. L. (2025). Ergothioneine Treatment Ameliorates the Pathological Phenotypes of Parkinson’s Disease Models. Journal of neurochemistry, 169(7), e70168.

Shukla, Y., Kulshrestha, O. P., & Khuteta, K. P. (1981). Ergothioneine content in normal and senile human cataractous lenses. The Indian journal of medical research, 73, 472–473.

Matsuda, Y., Ozawa, N., Shinozaki, T., Wakabayashi, K. I., Suzuki, K., Kawano, Y., Ohtsu, I., & Tatebayashi, Y. (2020). Ergothioneine, a metabolite of the gut bacterium Lactobacillus reuteri, protects against stress-induced sleep disturbances. Translational psychiatry, 10(1), 170.

Makoto Katsube a,*, Hiroshi Watanabe a, Kosuke Suzuki a, Takahiro Ishimoto b, Yoshitaka Tatebayashi c, Yukio Kato b, Norihito Murayama a Journal of Functional Foods 95 (2022) 105165 Food-derived antioxidant ergothioneine improves sleep difficulties in humans.

Smith E, Ottosson F, Hellstrand S, Ericson U, Orho-Melander M, Fernandez C, Melander O. Ergothioneine is associated with reduced mortality and decreased risk of cardiovascular disease. Heart. 2020 May;106(9):691–697.

Dare, A., Channa, M. L., & Nadar, A. (2021). L-ergothioneine and metformin alleviates liver injury in experimental type-2 diabetic rats via reduction of oxidative stress, inflammation, and hypertriglyceridemia. Canadian journal of physiology and pharmacology, 99(11), 1137–1147.

Tian, X., Cioccoloni, G., Sier, J. H., Naseem, K. M., Thorne, J. L., & Moore, J. B. (2021). Ergothioneine supplementation in people with metabolic syndrome (ErgMS): protocol for a randomised, double-blind, placebo-controlled pilot study. Pilot and feasibility studies, 7(1), 193.

Lu, Shelly C (2008). “Antioxidants in the treatment of chronic liver diseases: why is the efficacy evidence so weak in humans?.” Hepatology (Baltimore, Md.) vol. 48,5 : 1359–61.

Xiong K X, Guo H, Jia Y S, Liang Y Q, et al (2024). Supplement of food functional factor ergothioneine can effectively prevent liver injury in mice. Food Bioscience, Vol 57.

Ramachandraboopathi, S., Sathanantham, S. T. (2022). L-Ergothioneine protects hepatocytes against azathioprine-Induced toxicity via NIK/NF-?B signaling axis. International Journal of Health Sciences, 6(S7), 2233–2244.

Deiana, Monica et al. (2004). “L-ergothioneine modulates oxidative damage in the kidney and liver of rats in vivo: studies upon the profile of polyunsaturated fatty acids.” Clinical nutrition (Edinburgh, Scotland) vol. 23,2: 183–93.

Kimura, Chieko et al. (2005). “beta-Hydroxyergothioneine, a new ergothioneine derivative from the mushroom Lyophyllum connatum, and its protective activity against carbon tetrachloride-induced injury in primary culture hepatocytes.” Bioscience, biotechnology, and biochemistry vol. 69,2 : 357–63.

Bedirli, A., Sakrak, O., Muhtaroglu, S., Soyuer, I., Guler, I., Riza Erdogan, A., & Sozuer, E. M. (2004). Ergothioneine pretreatment protects the liver from ischemia-reperfusion injury caused by increasing hepatic heat shock protein 70. The Journal of surgical research, 122(1), 96–102.

Milito, Alfonsina, Mariarita Brancaccio, Giuseppe D’Argenio, and Immacolata Castellano. 2019. “Natural Sulfur-Containing Compounds: An Alternative Therapeutic Strategy against Liver Fibrosis” Cells 8, no. 11: 1356.

Mao, Y., Xie, Z., Zhang, X., Fu, Y., Yu, X., Deng, L., Zhang, X., Hou, B., Wang, X., Ma, M., & Ren, F. (2025). Ergothioneine Ameliorates Liver Fibrosis by Inhibiting Glycerophospholipids Metabolism and TGF-β /Smads Signaling Pathway: Based on Metabonomics and Network Pharmacology. Journal of applied toxicology : JAT, 45(3), 514–530.

Marone, P. A., Trampota, J., & Weisman, S. (2016). A Safety Evaluation of a Nature-Identical l-Ergothioneine in Sprague Dawley Rats. International journal of toxicology, 35(5), 568–583.

EFSA Panel on Dietetic Products, Nutrition and Allergies (NDA), Turck, D., Bresson, J. L., Burlingame, B., Dean, T., Fairweather-Tait, S., Heinonen, M., Hirsch-Ernst, K. I., Mangelsdorf, I., McArdle, H. J., Naska, A., Neuhäuser-Berthold, M., Nowicka, G., Pentieva, K., Sanz, Y., Siani, A., Sjödin, A., Stern, M., Tomé, D., Vinceti, M., van Loveren, H. (2017). Statement on the safety of synthetic l-ergothioneine as a novel food - supplementary dietary exposure and safety assessment for infants and young children, pregnant and breastfeeding women. EFSA journal. European Food Safety Authority, 15(11), e05060.

